# Pregnancy and Parenthood in Surgical Training

**DOI:** 10.1101/2023.04.19.23288808

**Authors:** Jessica Whitburn, Saiful Miah, Sarah A. Howles

## Abstract

**Objectives:** To describe pregnancy outcomes, rates of infertility, patterns of parental leave, and working schedules in surgical trainees in the United Kingdom.

**Design:** Cross sectional survey.

**Setting:** Surgical training programs in the United Kingdom

**Participants:** Four hundred and sixteen individuals who were enrolled on a surgical training program between June 2022 and March 2023.

**Main Outcome Measures:** Self-reported age, gender, infertility investigations, pregnancy loss, pregnancy-associated complications, live births, parental leave, and working patterns.

**Results:** Approximately half of all surgical trainees delayed attempting to have children due to training, over 80% regretted this decision and 23% of trainees had undergone fertility testing. Overall, childbearing surgical trainees experienced a pregnancy loss rate of 31%, and those aged less than 35 years had a pregnancy loss rate of 35%. A third of trainees did not take any time off work following pregnancy loss and over half of trainees did not disclose their loss to colleagues. Major pregnancy-associated complications occurred in 31% of pregnancies in surgical trainees, a significantly higher rate than pregnancies in a socio-demographically similar control group (9%, p=0.0001). Most trainees continued to work at night throughout their pregnancy and half continued to operate for more than 9 hours each week up until parental leave; trainees felt guilty for burdening their colleagues by reducing their workload. Childbearing surgical trainees on average took 10.2 months of parental leave whilst most non-childbearing surgical trainees took 2 weeks; two thirds of non-childbearing surgical trainees felt this was insufficient. After parental leave, 61% of childbearing and 15% of non-childbearing surgical trainees reduced their working hours to accommodate family life.

**Conclusion:** Surgical trainees often delay parenthood due to training and are at risk of high rates of infertility, pregnancy loss, and major pregnancy-associated complications. This study highlights the need for changes in surgical culture and training structures to improve obstetric health and facilitate family life for surgeons in training.

## Introduction

Women were first admitted to medical schools in the late 19^th^ century and since then, female representation in the medical profession has gradually increased^1^. By 2012, in the United Kingdom (UK) over 60% of doctors on the medical register under the age of 30 were women and currently the majority of medical students are female^2^. However, female representation within surgical specialities remains poor; in 2020 only 16% of all consultant surgeons in the UK were female, with variation by speciality from 7% of trauma and orthopaedic surgeons to 32% of paediatric surgeons^3^. Similarly, data from the United States indicate that in 2019 only 22% of all general surgeons were female^4^. Furthermore, surgical training programs have the greatest gender imbalance in England with women constituting only 30% of specialist surgical registrars and 27% of the surgical workforce as a whole^5^.

Workforce diversity is of benefit to society and is thought to improve multiple outcomes across organisations^6,7^. Thus, there has been a drive to increase understanding of the causes of this surgical gender gap and improve access for women to surgical careers. Studies have reported that surgery can be seen as an unattractive career option for many female undergraduates due to factors including perceived difficulties in maintaining family life, limited opportunities for flexible training, and a lack of role models^8–11^. Indeed, the Royal College of Surgeons of England commissioned Kennedy report of 2021 found that the experience of parents in surgery is *‘extremely challenging and stressful; it demands urgent attention’*. Furthermore, data from the USA and Ireland indicates that female surgeons have fewer children, are more likely to delay parenthood, are at greater risk of pregnancy-associated complications, and have higher rates of infertility^12–15^. It is unclear if these problems persist in other populations, and there is limited data from populations working in environments that aim to comply with the European Working Time Directive (EWTD), such as surgical trainees within the UK. It is probable that perceived difficulties in maintaining work-life balance in surgical careers and the reported risks to fertility and pregnancy contribute to gender imbalance in surgical training programs via impaired recruitment and retention.

We sought to increase understanding of experiences of parenthood in surgical training in the UK and define the impact of surgical working practices, which typically includes night shifts, manual handling, and long periods standing to operate, on fertility, and maternal and neonatal health.

## Methods

### Study design and data collection

We performed a descriptive cross-sectional survey in surgical trainees (residents) in the UK. Our methods are reported in accordance with the Checklist for Reporting Results of Internet Surveys^16^. A 192-item survey entitled ‘Pregnancy and Parenthood during Surgical training’ was generated (supplementary material). As no existing instrument was validated to evaluate our study aims, questions were developed based on previous surveys and literature review^12,15,17,18^. The resultant survey explores all aspects of pregnancy, childbearing, and parental leave, including information regarding level of training and working hours. Answers were recorded using a combination of binominal, multiple choice, and Likert scales. The survey was tested in male and female surgeons and iteratively revised. The web-based survey tool, JISC (2022, Bristol, UK), was used to administer the questionnaire, online completion was facilitated via a weblink.

The survey was distributed via nationally recognised/endorsed surgical specialty trainee groups and publicised on social media platforms. The survey was open for data collection from 24^th^ June – 24^th^ December 2022. All surgical trainees working in the UK were invited to participate to prevent bias. If a surgeon was a parent but not themselves childbearing, they were asked to answer questions regarding their partner’s pregnancies. These individuals served as a socio-demographically similar control group for pregnancy outcomes and are referred to as non-surgeon partners within the study. Male surgeons with female surgeon partners in training were excluded from analysis to prevent the same data being captured twice. Participation was voluntary and uncompensated; the participant information leaflet explained that survey completion would act as implied consent. Individual trainees were not identifiable from the survey. Ethical approval was gained via the Central University Research Ethics Committee, University of Oxford (Ref: R80778/RE001).

### Data analysis

Data were analysed in GraphPad Prism (version 9.5.0, California, USA). Significance testing was performed using χ_2_ or Fishers exact test for non-parametric binary data and Student t test for continuous variables. All tests of significance were 2-tailed. Missing data was omitted from analyses. Pregnancy loss rate was calculated using the formula: (total number of pregnancy losses/total number of pregnancies) x 100.

### Patient and public involvement

This study was designed and analysed by individuals who are, or have recently been, surgical trainees.

## Results

### Demographics

A total of 440 surgeons completed the survey and 416 surgeons (346 female and 70 male) were included in the final analysis (Supplemental Figure 1). The majority of participants were aged 30-39 years (Table 1) and responses spanned all training grades (5% CT1-2, 45% ST3-5, 35% ST6-8, 16% out of program/other). Most respondents were married or had a significant partner, childbearing respondents were more likely to be single, and white/caucasian (Table 1). Non childbearing surgeons were more likely to be in urological or ENT training, have a partner working <40 hrs per week, and have a partner working as a physician or surgeon (Table 1).

**Table 1:**
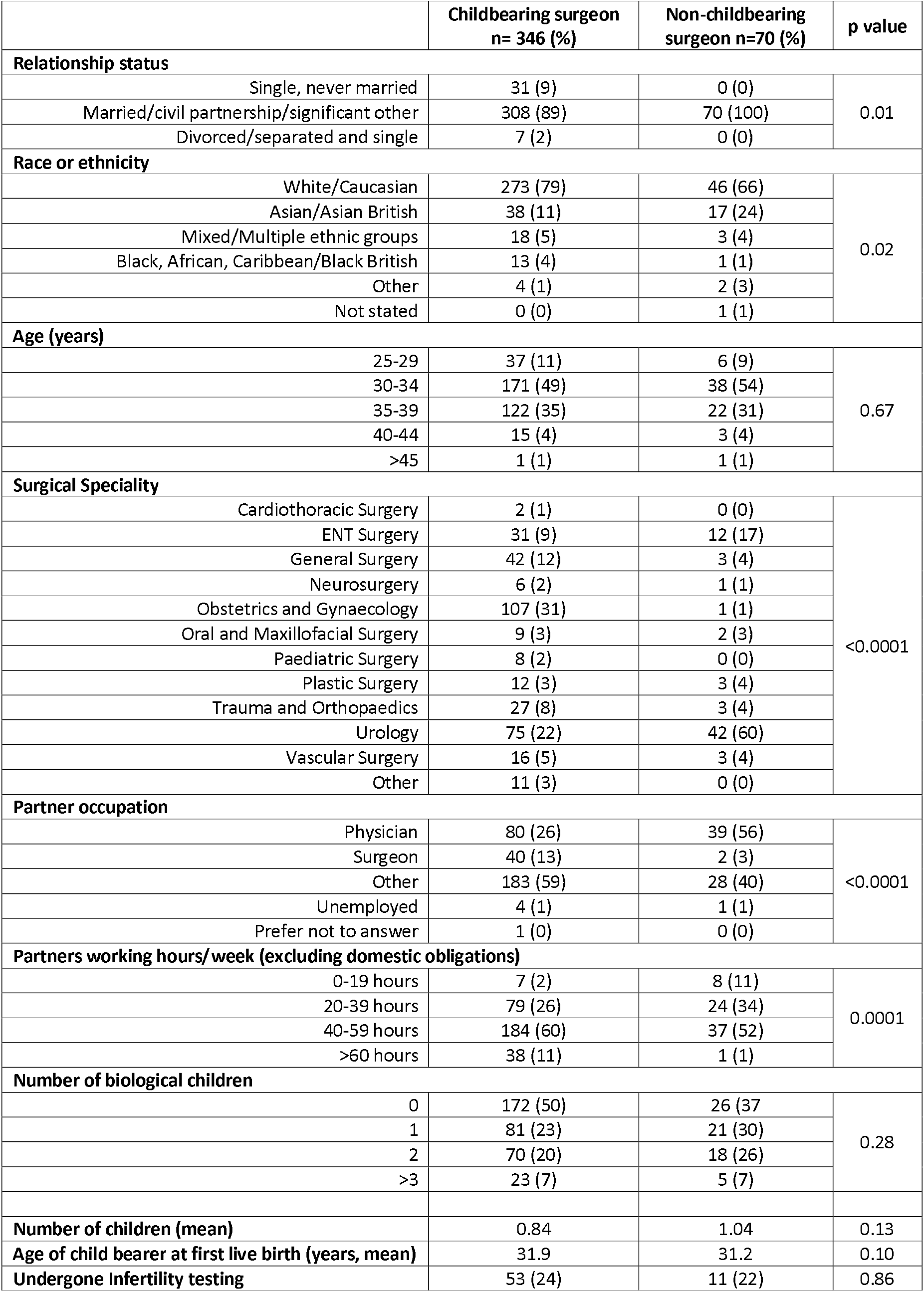
Characteristics of the survey participants.

### Childbearing

A total of 352 live births during surgical training were reported by 212 individuals (171 childbearing and 41 non-childbearing). Childbearing and non-childbearing trainee surgeons had similar numbers of children (Table 1). Fifty six percent of childbearing and 40% of non-childbearing surgical trainees delayed having children due to their training. A large proportion of those who delayed childbearing due to surgical training, regretted this decision (83% childbearing, 86% non-childbearing). Age of first pregnancy of childbearing surgeons and partners of non-childbearing surgeons was comparable (Table 1). Amongst the 402 respondents who were parents or had actively tried to conceive, 23% had undergone fertility testing (Table 1).

### Pregnancy loss and infertility

Of the 222 childbearing surgical trainees who had been pregnant, 36% had experienced a pregnancy loss and the pregnancy loss rate was 31%; these figures were comparable to data from non-surgeon partners (Table 2). In surgeons, in all age groups rates of pregnancy loss were higher than those reported in a large registry based study^19^. Furthermore, loss rates were 35% in those aged under 35 years at the time of survey completion, a rate which is over 3 times that which would be expected in this age group (Table 2). Data was not available regarding the ages of non-surgeon partners at time of pregnancy loss. The majority of pregnancy losses occurred before 12 weeks of gestation; no still births were reported (Table 2). After pregnancy loss, 1 in 3 childbearing surgical trainees did not take any time of work and only 47% disclosed their loss to a colleague. In general, surgical trainees who did share their loss with colleagues found them to be supportive (Supplemental Table 1).

**Table 2:**
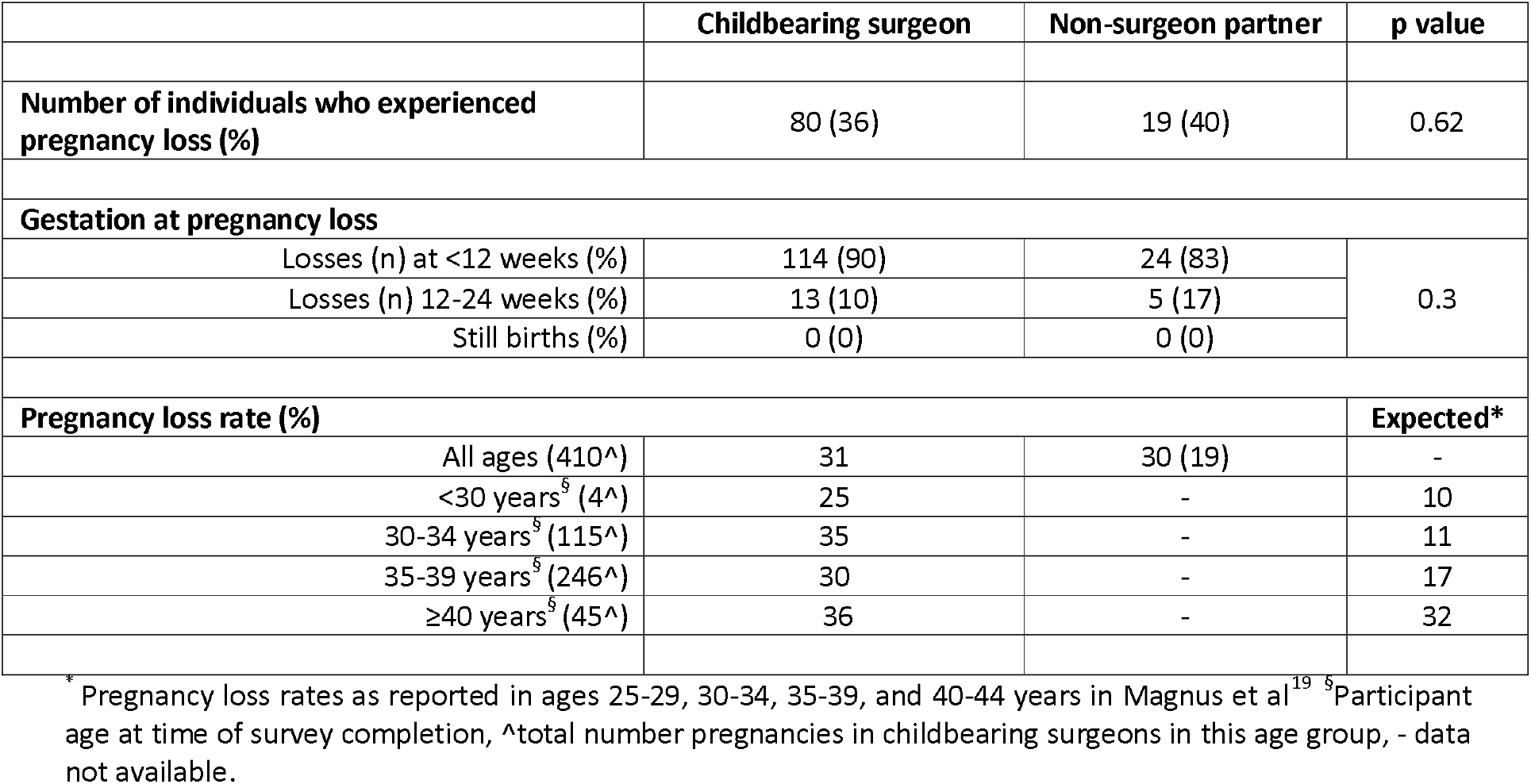
Pregnancy loss in surgical training.

### Pregnancy-associated complications

Almost 1 in 5 childbearing surgical trainees had a pregnancy complication that necessitated bedrest and nearly 1 in 10 needed more than 2 weeks off work, these figures are comparable to those of partners of surgical trainees (Table 3). The majority of trainees who required time away from work felt their colleagues and training programme were supportive of this time off (60% and 62%, respectively). Twelve childbearing surgical trainees reported a financial loss due to work restriction during pregnancy; financial losses ranged from £3,000 to £50,000 (median £7,500).

**Table 3:** Pregnancy complications in surgical training.

|  | Childbearing surgeon, n (%) | Non-surgeon partner, n (%) | p value |
| --- | --- | --- | --- |
| Pregnancy complications |  |  |  |
| Pregnancy with complication requiring bedrest | 50 (18) | 10 (14) | 0.6 |
| Pregnancy with one or more minor complication | 175 (62) | 35 (51) | 0.1 |
| Pregnancy with one or more major complication | 87 (31) | 6 (9) | 0.0001 |
| Delivery with one or more intrapartum complication | 60 (21) | 9 (13) | 0.13 |
| Delivery with one or more postpartum complication | 27 (10) | 4 (6) | 0.48 |
| Neonatal complications |  |  |  |
| NICU stay | 17 (6) | 3 (4) | 0.74 |
| Delivery <38 weeks | 35 (12) | 7 (10) |  |
| Birth weight |  |  |  |
| <2.5kg | 10 (4) | 6 (8) | 0.21 |
| 2.5-3kg | 54 (19) | 15 (22) |  |
| 3-4kg | 188 (66) | 41 (59) |  |
| >4kg | 30 (11) | 5 (7) |  |
| No answer | 1 (0) | 2 (3) |  |

Minor pregnancy-associated complications included hyperemesis, musculoskeletal issues, gestational diabetes, and gastro-oesophageal reflux. Trainees experienced minor complications in 175 pregnancies out of a total of 282 pregnancies (62%), a proportion not significantly different to rates of minor pregnancy-associated complications in the partners of non-childbearing surgical trainees (Table 3). Major pregnancy-associated complications were defined as pre-eclampsia or hypertension, placental abruption or bleeding in pregnancy, placenta praevia or accrete, intrauterine growth restriction (IUGR), and placental insufficiency (including oligohydramnios). Childbearing surgical trainees were significantly more likely to experience major pregnancy complications than the partners of non-childbearing surgical trainees (Table 3). Rates of neonatal, intrapartum, and postpartum complications were higher in childbearing surgical trainees than in the partners of surgical trainees, however statistical significance was not reached in this small sample size (Table 3). Babies of childbearing surgical trainees were comparable in weight to those of non-childbearing surgical trainees (Table 3).

### Work schedule during pregnancy

Over 70% of childbearing surgical trainees worked for 40 hours or more each week during pregnancy and 4% worked more than 60 hours per week. Most trainees continued to work at night throughout their pregnancy and half continued to operate for more than 9 hours each week up until maternity leave (Table 4). The majority (70%) of childbearing surgical trainees altered their work schedule during pregnancy; 77% felt guilty for burdening their colleagues by reducing their workload. Of those trainees who did not alter their work schedule in pregnancy, 40% made this decision to avoid being ‘considered weak’ and 35% because of concerns surrounding burdening colleagues.

**Table 4:** Working patterns during pregnancy.

|  | Childbearing surgeon, n (%) | Non-surgeon partner, n (%) | p value |
| --- | --- | --- | --- |
| Average hours worked per week |  |  |  |
| <40 | 78 (28) | 33 (48) | 0.003 |
| 40-60 | 190 (67) | 36 (52) |  |
| >60 | 10 (4) | 0 |  |
| No answer | 5 (2) | 0 |  |
| Frequency of overnight on call/nightshifts |  |  |  |
| None | 53 (19) |  |  |
| 2-4/month | 147 (52) |  |  |
| 4-6/month | 66 (23) |  |  |
| >6/month | 15 (5) |  |  |
| No answer | 2 (1) |  |  |
| Hours operating last trimester |  |  |  |
| 0-8 | 140 (49) |  |  |
| 9-12 | 75 (27) |  |  |
| 13-16 | 41 (14) |  |  |
| >16 | 26 (9) |  |  |
| No answer | 1 (0.4) |  |  |
| Average hours worked per week in last trimester |  |  |  |
| <30 | 78 (28) |  |  |
| 30-40 | 96 (34) |  |  |
| 41-60 | 105 (37) |  |  |
| >60 | 2 (1) |  |  |
| No answer | 2 (1) |  |  |

### Breastfeeding

The majority (98%) of childbearing surgical trainees wanted to breastfeed their child, and 85% achieved this for over 6 months. Over two thirds of surgical trainees (79%) breastfed for as long as they wanted to. Surgical trainees who stopped breastfeeding early cited inadequate work provisions to enable continued breastfeeding as the most common reason; other barriers included insufficient milk supply, struggles with work schedules and fatigue or illness.

### Parental leave

Childbearing surgical trainees took a mean of 10.2 months of parental leave, which was comparable to parental leave taken by partners of non-childbearing surgical trainees (10.1 months). The majority of non-childbearing surgical trainees took 2 weeks of parental leave (n=30, 73%); only 3 respondents took more than 8 weeks of leave, and 2 did not take any leave. Two thirds of non-childbearing surgical trainees felt that the amount of leave taken was insufficient (n=27, 66%) but cited concerns that taking more time would be viewed negatively by colleagues (33%) and that they were reluctant to prolong their training (26%). The non-childbearing surgical trainees who took more than 8 weeks parental leave did not find this easy to arrange. Only a third of non-childbearing surgical trainees found they were able to organise their parental leave without any difficulties (Supplemental Table 2).

### Return to work

On return to work after parental leave, 61% (n=104) of childbearing surgical trainees reduced their working hours to less than 40 hours per week, compared to only 15% (n=6) of non-childbearing surgical trainees. Approximately 10% of both childbearing and non-childbearing surgical trainees altered their work schedules after parental leave to accommodate family life but continued to work more than 40 hours per week. Almost 1 in 4 non-childbearing trainees were unhappy with their work schedule on return to work but did not change it, the main factor preventing non-childbearing trainees from adjusting their work schedule was worry that it would be viewed negatively by colleagues (n=5, 56%, Supplemental Table 3).

## Discussion

This study of pregnancy and parenthood in 416 UK surgical trainees reveals that surgeons in training under 35 years of age experience a pregnancy loss rate of 35%, a rate that is more than three times that which would be expected in women of a similar age. In childbearing surgeons of all ages, rates of pregnancy loss are ∼30%, which is comparable to loss rates reported previously in pregnant female surgeons^17,19^. Our data revealed similar pregnancy loss rates in the childbearing partners of surgical trainees; this may reflect the fact that nearly 60% of childbearing partners of surgical trainees in this survey were doctors, investigation of pregnancy loss rates in other health care professionals should be undertaken. Previous studies have reported that night shifts and working more than 40 hours a week are associated with higher risks of miscarriage^20–22^. Whilst working patterns prior to miscarriage were not assessed in our study, we report that 57% of childbearing surgical trainees worked >40 hours per week during pregnancies that resulted in a live birth and that 81% continuing to work night shifts or take part in an overnight on call during pregnancy. Limiting working hours and overnight commitments for pregnant surgical trainees may help reduce miscarriage rates.

The psychological impact of early pregnancy loss is well recognised^23–25^. However, 33% of childbearing surgical trainees in this study did not take any time off work after miscarriage and approximately half did not reveal to colleagues that they had experienced a pregnancy loss. A survey undertaken in the UK found that 27% of surgical trainees did not feel supported by their department during pregnancy, this may explain hesitancy to disclose information relating to pregnancy loss^26^. It is well established that doctors are less likely to take days off sick in comparison to other healthcare workers with many citing that taking time off will negatively affect their careers and burden their colleagues with extra workload^27,28^. Surgical training programs should consider offering bereavement support and mandated leave after pregnancy loss to reduce this burden of guilt. Appointment of a lead clinician for pregnancy support may help trainees navigate this difficult area, providing a single point of contact to facilitate changes to working schedules in early pregnancy and provide guidance after loss.

Studies have previously reported that female physicians and surgeons commonly delay pregnancy due to training commitments despite well-recognised increases infertility and obstetric risks after 35 years of age^15^. Our survey reveals that approximately half of all surgical trainees in the UK delay childbearing and that the mean age at birth of first child amongst childbearing trainees is 31.9 years. In the UK population, the mean age at birth of first child is 27.8 years^29^. The vast majority (>80%) of trainee surgeons regretted their decision to delay childbearing, and we found that both male and female surgeons underwent infertility investigations more frequently than would be expected in the general UK population^30^. This data is in keeping with data from the USA which reported higher rates of infertility problems and use of assisted reproduction technologies amongst female orthopaedic surgeons and postgraduate medical trainees, but is in contrast to a study in Irish surgical trainees^13,15,31^. Cultural change and open conversations may enable surgical trainees to make better informed decisions about timing of attempts to become parents so that fewer individuals regret decisions to postpone pregnancy.

Our study indicates that major complications occur in ∼30% of pregnancies in childbearing surgical trainees, a rate that was significantly higher than a socio-demographically matched control group and in keeping with other studies of pregnancy in childbearing surgeons^12,13,15,17^. The causes of these high pregnancy complication rates cannot be elucidated by this survey-study, however, associations of adverse pregnancy outcomes with working more than 40 hours a week, prolonged standing, high fatigue scores, shift and night work have been reported; these factors are common in surgical training^20,22,32,33^. Furthermore, evidence exists that surgical trainees of both sexes omit healthcare maintenance activities including medical check-ups and exercise^34^. Our study suggests that even in combination with statutory maternity leave policies, the rest and working time requirements of the EWTD affords insufficient protection to pregnant surgical trainees. In view of the significantly increased risk of major pregnancy complication in this group it has previously been recommended that childbearing surgeons should be treated as a high-risk obstetric group, a recommendation that is supported by our data^13,35^. Furthermore, our study suggests that employers should, as a matter of priority, offer altered working schedules to pregnant surgical trainees to protect obstetric health.

Our study reveals that financial losses due to work restrictions in pregnancy are not uncommon for surgical trainees, steps should be taken to address this source of gender pay gap.

The WHO recommends that infants are exclusively breastfed for the first six months of life, and that mothers should be encouraged to breastfeed for at least a year to provide protection from illnesses and long-term diseases. The rate of initiated and continued breastfeeding in our study was higher than that reported amongst US resident physicians and orthopaedic surgeons and the UK national average^13,36,37^. Over 75% of surgical trainees breastfed for as long as they wanted, however, among those who stopped early, 35% reported inadequate work provisions as a factor in early cessation.

All UK employees have the right to 52 weeks of maternity leave, and most childbearing individuals will qualify for 39 weeks of statutory maternity pay. In our study, childbearing surgical trainees took an average of 10.2 months of parental leave, the majority of non-childbearing surgical trainees took 1-2 weeks of parental leave. Most non-childbearing surgical trainees felt that their parental leave was insufficient but were concerned that taking more might be viewed negatively by colleagues. Furthermore, only 15% of non-childbearing trainees reduced their working hours after parental leave compared to 61% of childbearing trainees. It may be that adopting a Scandinavian model of shared paid parental leave, with periods ringfenced for each parent, would lead to a more equitable approach to childcare in the medical workforce.

We predict that selection bias was present in this study and that non-childbearing trainees and those who experienced uncomplicated pregnancies were less likely to respond. Indeed, most survey responses were from female trainees despite male trainees comprising most of the surgical workforce. We also recognise that our findings are limited to experiences of surgical trainees in the UK, and therefore caution should be exercised before initiating changes to working practices outside of this setting.

In summary, this study provides a contemporary picture of the risks and challenges facing surgical trainees in the UK who are, or wish to become, parents. Our data highlights the need for urgent action to reduce rates of miscarriage and major pregnancy complications in EWTD compliant settings. It is likely that altering working schedules and limiting physical exertion will mitigate these risks, however, consideration will need to be taken of the impact of changes in working patterns on surgical training. Furthermore, this study indicates that cultural change is needed to empower pregnant trainees to ask for working modifications and time off in pregnancy and to facilitate the ability of non-childbearing surgical parents to take longer periods of parental leave and to access alterations in work schedules. Training programs and national surgical associations must evolve to progress diversity in the surgical workforce and protect the obstetric health of surgeons.

## Data Availability

All data produced in the present study are available upon reasonable request to the authors

## Acknowledgements

We acknowledge the contribution of the surgeons in training who provided data for this study.

## Data sharing statement

Data utilised in the preparation of this manuscript will be made available to researchers upon reasonable request to the corresponding author.

## Ethical approval

This research was conducted under ethical approval granted via the Central University Research Ethics Committee, University of Oxford (Ref: R80778/RE001).

## Transparency statement

The manuscript’s guarantors (J.W. and S.A.H.) affirm that the manuscript is an honest, accurate, and transparent account of the study being reported; that no important aspects of the study have been omitted; and that any discrepancies from the study as planned (and, if relevant, registered) have been explained.

## Contributors

J.W., S.M., and S.A.H designed this study. J.W. and S.A.H. analysed and interpreted data. J.W. and S.A.H. wrote the first draft of the manuscript. All authors participated in the preparation of the manuscript by reading and commenting on the draft prior to submission. The corresponding author attests that all listed authors meet authorship criteria and that no others meeting the criteria have been omitted.

## Copyright statement

The Corresponding Author has the right to grant on behalf of all authors and does grant on behalf of all authors, a worldwide licence to the Publishers and its licensees in perpetuity, in all forms, formats and media (whether known now or created in the future), to i) publish, reproduce, distribute, display and store the Contribution, ii) translate the Contribution into other languages, create adaptations, reprints, include within collections and create summaries, extracts and/or, abstracts of the Contribution, iii) create any other derivative work(s) based on the Contribution, iv) to exploit all subsidiary rights in the Contribution, v) the inclusion of electronic links from the Contribution to third party material where-ever it may be located; and, vi) licence any third party to do any or all of the above.

