## Supplemental for "Pregnancy and Parenthood in Surgical Training"

**This appendix has been provided by the authors to give additional information about their work**

**Supplemental Figure 1: Flow chart of survey participants and exclusions**

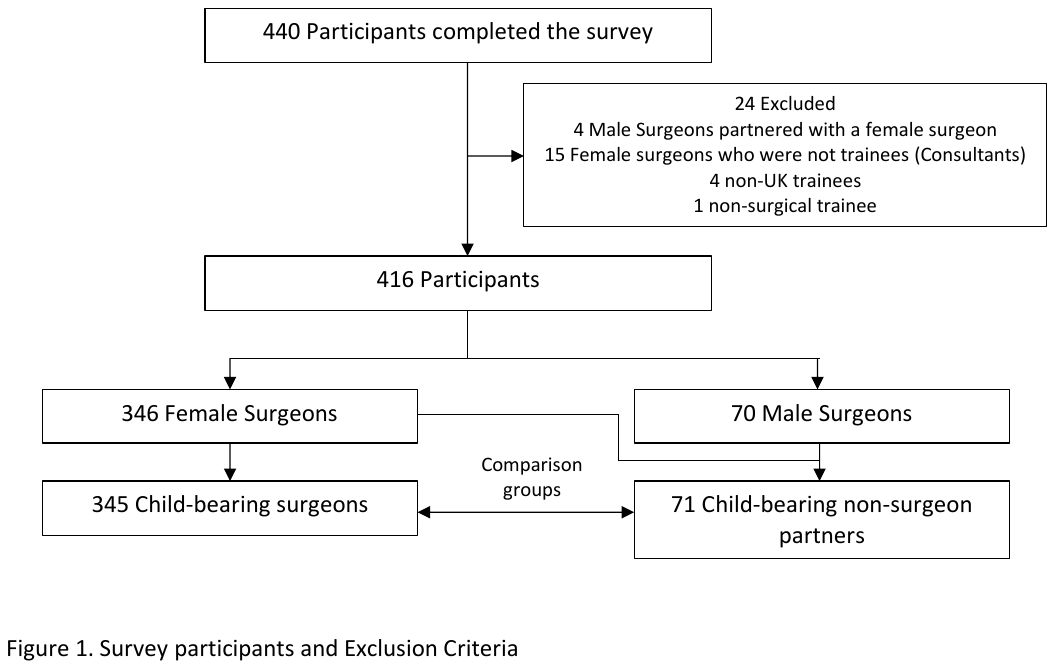

**Supplemental Table 1: Time off after pregnancy loss**

|  | **Childbearing surgeon, n (%)** |
| --- | --- |
| **Time off after pregnancy loss** | |
| No time off | 29 (33) |
| 1-7 days | 37 (43) |
| 1-2 weeks | 14 (16) |
| 2-3 weeks | 0 (0) |
| 3-4 weeks | 5 (6) |
| >4 weeks | 2 (2) |
| **I felt my colleagues were supportive of time off after pregnancy loss** | |
| Strongly agree | 8 (9) |
| Agree | 24 (28) |
| I didn’t tell them | 46 (53) |
| Disagree | 6 (7) |
| Strongly disagree | 3 (3) |
| **I felt my supervisors/training programme director were supportive of time off after pregnancy loss** | |
| Strongly agree | 14 (16) |
| Agree | 22 (25) |
| I didn’t tell them | 41 (47) |
| Disagree | 7 (8) |
| Strongly disagree | 3 (3) |

**Supplemental Table 2: Parental leave for non-childbearing surgical trainees**

|  | **Non-childbearing surgeon, n (%)** |
| --- | --- |
| **Parental leave (weeks)** | |
| None | 2 (5) |
| 1-2 | 31 (76) |
| 3-8 | 4 (10) |
| >8 | 3 (7) |
| No answer | 1 (2) |
| **Was it easy to arrange parental leave?** | |
| Yes, no difficulties | 15 (37) |
| Some difficulties but organised to my satisfaction | 13 (32) |
| No, it was not easy to arrange | 9 (22) |
| N/A | 3 (7) |
| No answer | 1 (2) |

**Supplemental Table 3: Return to work after parental leave**

|  | **Childbearing surgeon, n (%)** | **Non-childbearing surgeon, n (%)** |
| --- | --- | --- |
| **Changes to work pattern on return to work** | | |
| No change, happy with work schedule | 29 (17) | 21 (51) |
| No change, unhappy with work schedule | 17 (10) | 9 (22) |
| Altered work schedule, ≥ 40 hours/week | 18 (11) | 4 (10) |
| Altered work schedule, <40 hours/week | 104 (61) | 6 (15) |
| No answer | 3 (2) | 1 (2) |
| **If working <40 hours, what % worked** | | |
| 50% | 3 (3) | 1 (17) |
| 60% | 40 (38) | 1 (17) |
| 70% | 6 (6) | 0 (0) |
| 80% | 47 (45) | 4 (67) |
| 90% | 5 (5) | 0 (0) |
| Not confirmed | 1 (1) | 0 (0) |
| No answer | 1 (2) | 0 (0) |
